# Combining models to generate a consensus effective reproduction number *R* for the COVID-19 epidemic status in England

**DOI:** 10.1101/2023.02.27.23286501

**Authors:** Harrison Manley, Josie Park, Luke Bevan, Alberto Sanchez-Marroquin, Gabriel Danelian, Thomas Bayley, Veronica Bowman, Thomas Maishman, Thomas Finnie, André Charlett, Nicholas A Watkins, Johanna Hutchinson, Steven Riley, Nowcasts Model Contributing Group, Jasmina Panovska-Griffiths

## Abstract

The effective reproduction number *R* was widely accepted as a key indicator during the early stages of the COVID-19 pandemic. In the UK, the *R* value published on the UK Government Dashboard has been generated as a combined value from an ensemble of epidemiological models via a collaborative initiative between academia and government. In this paper we outline this collaborative modelling approach and illustrate how, by using an established combination method, a combined *R* estimate can be generated from an ensemble of epidemiological models. We analyse the *R* values calculated for the period between April 2021 and December 2021, to show that this *R* is robust to different model weighting methods and ensemble size, and that using heterogeneous data sources for validation increases its robustness and reduces the biases and limitations associated with a single source of data. We discuss how *R* can be generated from different data sources and is therefore a good summary indicator of the current dynamics in an epidemic.

## 1 Introduction

Since the onset of the coronavirus disease in early 2020 (COVID-19) as a pandemic, mathematical modelling has been widely used to generate policy-relevant evidence. Mathematical modelling provides a framework for simulating the dynamics of the pandemic. When parameterised with, and calibrated to data, this can be used to generate projections of future epidemic trajectories as well as to track the current epidemic status. Epidemiological estimates such as the reproduction number *R* derived from models can be useful tools for such epidemic status tracking.

The reproduction number *R* is a measure of the infectious potential of a disease and represents the average number of secondary infections that emerge from one infection [32]. At the onset of a new disease, in a naive, fully susceptible population, the basic reproduction number *R*_0_ represents the average number of secondary infections stemming from an initial case. In contrast to *R*_0_, *R* is the reproduction number at any time during an epidemic - often referred to as the *effective reproduction number R*_*e*_ or *temporal reproduction number R*_*t*_ [5]. It reflects the average number of secondary infections generated from a population consisting of susceptible, exposed and immune individuals, and potential changes in mixing and the presence of interventions.

The growth rate *r* represents the rate at which the epidemic is growing during the exponential phase of epidemic growth. In epidemiological modelling, *r* and *R* are related via the generation time (*τ* ) of the epidemic [5]. Mathematically, this is expressed as

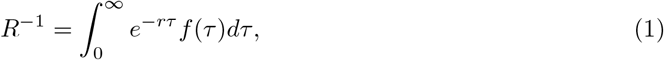

where *τ* is the time since infection, and *f* (*τ* ) is the probability density function for the time of infection, or the generation time distribution. As evident from this equation, changes in *τ, r* and *R* affect each other. Specifically, for fixed *τ, r* and *R* increase/decrease in tandem; for fixed *r, τ* and *R* increase/decrease in tandem while for fixed *R*, increasing *τ* means decreasing *r* and vice versa.

While *R* is reflective of the current strength of transmission, *r* is reflective of the transmission speed [20]. Both provide information about the impact of control measures. For example, if an intervention is imposed and *R* is consequentially reduced to below the *R* = 1 threshold, or *r* is reduced to below 0 threshold, this suggests that the intervention has had an impact on reducing onward transmission. However, when providing policy advice during the COVID-19 epidemic, *R* was used as it is more easily interpretable than *r* and does not require a conceptual understanding of exponential growth or decay, so is therefore simpler to explain to the public. Additionally, *R* at the onset of the epidemic (*R*_0_) provides information on the likely level of herd immunity necessary. In a homogeneous population, the herd immunity threshold as a percentage of the population, *I*_*c*_, can be calculated as:

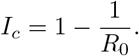

Which suggests that the more people that become infected by each individual who has the virus, the higher the proportion of the population that needs to be immune to reach herd immunity [23]. Although it should be noted that this is subject to large uncertainties due to the difficulty in calculating *R*_0_, which leads to differing estimates of *I*_*c*_, and should therefore be used with care [30, 17]. Further details on *R* and the differing methodologies for calculating the reproduction number can be found in Section 2.1.

In the UK, the Scientific Advisory Group for Emergencies (SAGE), is activated in response to emergencies and is made up of several sub-groups consisting of experts relating to different scientific fields [25]. These sub-groups are often called upon in order to provide evidence to the UK government relating to key policy questions. One of these groups is the Scientific Pandemic Influenza Group on Modelling - Operational (SPI-M-O) which has been leading the modelling of the COVID-19 epidemic since its onset [10]. SPI-M-O primarily consists of experts in infectious diseases modelling.

In early 2021, a formal collaboration between SPI-M-O and the UK Health Security Agency Epidemiological Ensemble modelling (UKHSA Epi-Ensemble) group was established, which has provided the UK government with weekly estimates of key epidemiological indicators, including the effective reproduction number *R* [26] throughout 2021-2023. The consensus values were generated as a combined estimate from a set of epidemiological models maintained and run by members of SPI-M-O and the UKHSA Epi-Ensemble, and were combined using a random effects meta analysis approach with equal weighting applied [42], with visualisation implemented using CrystalCast developed by the Defence Science and Technology Laboratory (DSTL)[49]. The combined estimates were agreed in a weekly meeting of the UKHSA Epidemiology Modelling Review Group (EMRG), attended by government modellers and policy stakeholders as well as academic modellers.

Generating a combined ensemble estimate in place of a single model truth can lead to improved predictive power [54], allows an increased robustness of the outcomes, and is a useful tool for policy makers [47]. Generating a combined estimate from a set of models is not a new concept; they are widely used across many disciplines; in forecasting the weather [41], hydrology [31], flood losses [22], in cancer prediction [59] and climate modelling [43]. Within infectious diseases, combined model estimates have been applied to modelling HIV [21], influenza [48] and Ebola [51, 13] transmission, and recently for outbreak analysis related to COVID-19 in the USA [53] and Europe [9].

While mathematical models have been used to offer informed advice to the scientific community and policy makers throughout the COVID-19 pandemic across a number of countries, the use of modelling has differed. For example, modellers in the United States, in conjunction with the Center for Disease Control (CDC), published ensemble forecasts using a wide variety of mathematical models [16, 47]. These models had focused on forecasting new cases, hospitalisations and deaths at a national and state-level but did not estimate *R* or *r* specifically. On the other hand, in New Zealand and Italy modellers advising the government have compared estimates of *R* obtained from different models but without producing formal combined estimates [36, 15]. In Norway, multiple data sources including confirmed cases, proportion of COVID-19 attributable hospital admissions and a national symptom survey were used to estimate *r* over the course of the pandemic, but only one model has been used to estimate *R* from these sources [45]. Similarly, the Robert Koch Institute in Germany only used a single model to estimate *R* which depended on nowcasting estimates of the number of new cases [50].

As noted above, in the UK, since the onset of the pandemic a set of mathematical models developed, maintained and applied by the members of SPI-M-O and the UKHSA Epi-Ensemble have been used to track epidemic status, including generating *R* and *r* alongside estimates of incidence and prevalence. The *R* value published on the UK Government Dashboard [33] has been generated as a combined value from these models and agreed at the weekly EMRG meeting.

The value in getting a combined value from across models and datasets is not just in the averaging of those estimates with weighting, but also in the formation of a community that are constantly discussing the outcomes, the assumptions, the input data identifying the drivers behind the differences across models. This is especially important when generating *R*. While doubling time and *r* can be thought of as almost features of the data, requiring very few assumptions, the move to *R* requires a set of subjective assumptions. This is why there is a need to have multiple groups making different assumptions leading to heterogeneous outcomes which can be discussed, understood and combined. When *R* can be generated using different data sets, in addition to different models, this is particularly important. The development of the formal collaboration between the modellers at UKHSA Epi-Ensemble and within SPI-M-O, and the weekly technical meetings of the group and the follow up EMRG meeting, gave a platform for informed discussions of the similarities and the differences across models’ nowcast estimates, and provided a place where decisions could be made on whether to include or exclude a given model from the combined estimate.

This paper outlines the process of this collaboration between government and academia to continually generate estimates for the effective reproduction number in England over the COVID-19 epidemic. Specifically, we outline how a previously established combination method, described in [42], has been applied in the UK throughout the COVID-19 pandemic. We detail our approach of generating a consensus value of *R* from an ensemble of epidemiological models applied to the English epidemic. We illustrate the process, show how a combined *R* estimate has been generated in April 2021 and in September 2021 and explore the robustness of the combined *R* value on the size and weighting of the models’ combination. By comparing the change in *R* with the change in measurable data COVID-19 cases, hospitalisations and deaths, we also explore whether *R* can be a good indicator of epidemic status.

## 2 Methodology

### 2.1 Outline of epidemiological models used to produce *R* values

Generating an *R* estimate requires a model of some kind with subjective assumptions and information from other sources. Our modelling ensemble comprised mathematical models that were developed, adapted and used throughout 2020-2022, to model the COVID-19 epidemic in England; to generate epidemic metrics such as *R, r*, incidence and prevalence; and to produce medium term projections (MTPs) of hospital admissions, hospital bed occupancy and deaths. The MTPs will be explored separately in a future publication. These models fall into three broad groups, as described in [18] and [5]: population based models (PBMs), data-driven models (DDMs) and agent-based models (ABMs). The models in the ensemble can be split further into three broad categories based on the data they primarily used to inform their estimates: case-based models, admissions-based models, and models that were fitted to both case data and hospital data. For the purposes of this study, models that were fitted to survey data are categorised as case-based models as they were focused on detecting incidence of the disease, though there were differing delays associated with models that were fitted to cases and models that were fitted to survey data. There are drawbacks and advantages associated with fitting to either cases or admissions. Case data is highly sensitive to ascertainment biases. For example, an under ascertainment of cases may be related to weekend/weekdays periods, with people with milder symptoms over the weekend less likely to get confirmed than during the weekdays. The scale of these biases have varied greatly over time. Therefore, models that were fitted to case counts or positivity must be interpreted in the context of testing behaviours and policies at the time. However, admissions data is not free from bias either, as they depend on input from physicians and other hospital staff, which means that weekend/weekday effects are likely. In addition, the likelihood of being admitted to hospital varies greatly by age. Hence, without age-stratification in the model, it is likely that community transmission is under-estimated among younger age groups. Furthermore, the delay between being infected with COVID-19 and being admitted to hospital is on average far greater than that between infection and receiving a positive test. This presented difficulties when trying to produce timely estimates of community transmission. Table 1 lists these models along with the type of data they were fitted to, and whether or not they were run internally by either the UKHSA Epi-Ensemble or a Devolved Administration (DA) department.

**Table 1.**
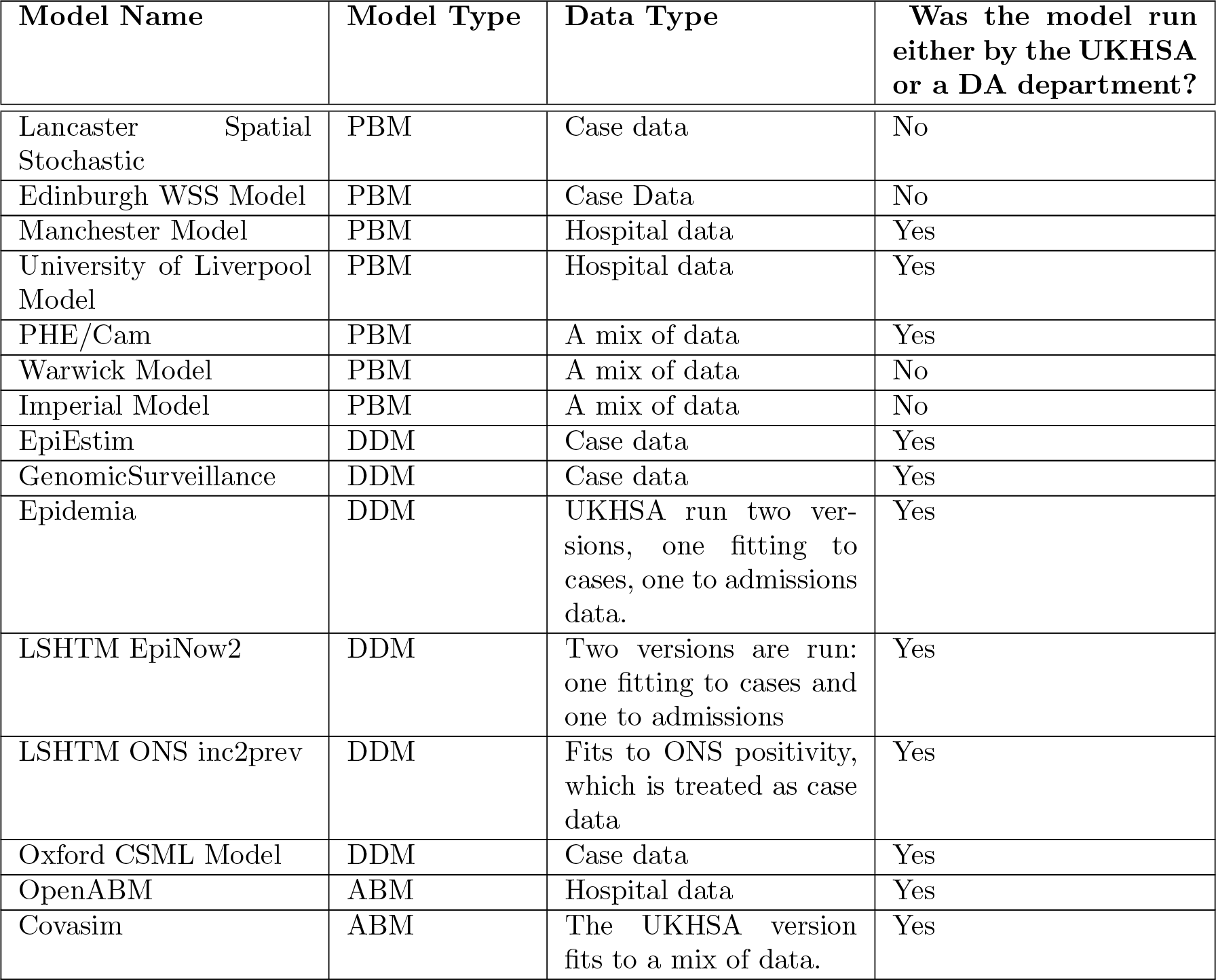
Table detailing the UKHSA/SPI-M-O models split by model type and the data to which they fit to.

While these models can be broadly stratified into the PBM, DDM and ABM groups, each model within the group has distinctive characteristics. For example, EpiEstim followed the methodology described in [14], and therefore assumed a consistent relationship between infections and cases. The estimated *R* was therefore only robust when the ascertainment rate was roughly constant. While GenSur shared this same limitation, Epidemia and OxfordCSML did not make this assumption [24]. Furthermore, renewal equation based models tend to be semi-mechanistic i.e. assuming that the effects of interventions are absorbed into the data to which they fit. In contrast, fully mechanistic models such as the SEIR population-based and the agent-based models, explicitly modelled the effects of interventions such as Test-Trace-Isolate strategies and imposing and removing of social distancing measures.

In epidemiological models, the structure of the model determines the method to calculate *R* and depends on the assumptions and data sets used to parameterise and validate the model [56].

In the classic compartmental Susceptible-Exposed-Infected-Recovered (SEIR) model, *R*_0_ = *β * c/γ* where *β* is the transmission probability, *c* is the number of contacts *c* and 1*/γ* is the infectiousness period (average time that an individual is infectious for). *R* is typically calculated from SEIR models as the largest eigenvalue of the next generation matrix (NGM), which can be expressed as *FV* ^*−*1^, where *F* represents infection rates, and *V* recovery rates [19, 12]. *R* can also be estimated using the renewal equation [29, 34]:

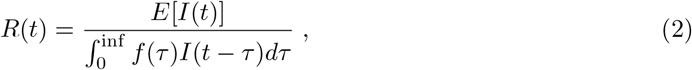

where *I*_*t*_ is number of infections as time *t*, and *E*[] denotes the expected value.

Across the dynamical models that comprise sets of differential equations in our ensemble, such as the Manchester or the University of Liverpool models, *R* was estimated by inferring the rate of transmission within the model, which was fitted to observed data on cases, hospitalisations, deaths or their combination. Some more complex dynamical models, such as PHE/Cambridge model or the Imperial (sircovid) model, explicitly calculated R as the largest eigenvalue of the NGM.

There is also a difference in how R was estimated between compartmental and agent- or individual-based models. In agent-based models, such as Covasim, it is possible simply to count exactly how many secondary infections are caused by each primary infection at any stage of the epidemic and hence explicitly calculate *R*.

A third approach, and characteristic of the data-driven models in our ensemble, used statistical models to estimate R empirically from the notification data. These methods made minimal structural assumptions about epidemic dynamics, and only required users to specify the distribution of the generation interval. A selection of models in this category in the ensemble were formulated based on Equation (1). For example, where the generation time is described by a gamma distribution, with shape *a* and rate *b, R* can be expressed in terms of the growth rate *r* as:

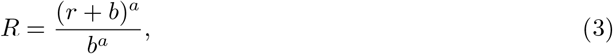

A high-level description of the methods used to calculate *R*, along with an outline of the main characteristics of each model are given in Table A1 in the appendix

### 2.2 Combining model estimates to generate a consensus *R*

To generate combined *R* estimates from the ensemble of models, we used the statistical model developed as a collaboration between DSTL, University of Southampton and University of Liverpool with the underlying methodology described in [42]. We present a high-level outline of the method below. Each of the epidemiological models described in Table A1 and calibrated to the data as outlined in Table 1, generated 5th, 25th, 50th, 75th and 95th percentile estimates for *R*. Using these, a mean and a standard deviation for each model’s *R* estimate was generated. The mean of the *i*^*th*^ model, *y*_*i*_, was initially estimated as the median (or 50^*th*^ quantile), and the standard deviation was calculated as follows:

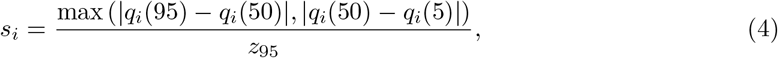

Where *q*_*i*_(*x*) represented the *x*^*th*^ quantile of the *i*^*th*^ model, and *z*_95_ is the z-score for the 90% confidence interval (CI) of the standard normal distribution. Where model estimates were highly skewed, a skewness correction calculation was applied to provide alternative estimates for the mean and standard error (see [42] for further details). Otherwise, the distribution of the model estimates for *R* were assumed to be symmetric.

These estimates were then combined using a random effects model which allowed for differences in model structure and did not assume that models shared a common effect size. The random effects statistical model was described by:

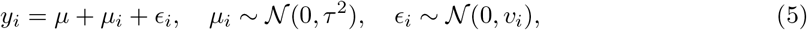

where the estimated mean for model *i* is denoted by *y*_*i*_ and standard error denoted by 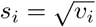. The model was fitted to provide estimates for *μ* and *τ* which are the mean and standard deviation of the true effect size respectively. The between model variance, *τ* ^2^, was estimated using the restricted maximum likelihood method, and the confidence intervals of the mean true overall effect size are estimated using the standard Wald-type method. The models were equally weighted (see next section for more details) and the range of *R* was rounded out to one decimal place, by using the lower and upper bounds respectively. Further details of other methods used for calculating the between group errors and CIs are provided in [57].

### 2.3 Collaborating across government and academia to produce a consensus nowcast

The process of cross academic and government to generate consensus *R* was done in several steps. Firtsly, the outputs from the models detailed in Table A1 were submitted by the modellers to the UKHSA Epi-Ensemble team weekly. The team then combined the model estimates using CrystalCast to generate a combined estimate for *R, r*, incidence and prevalence in England, the English regions and the Devolved Administrations. The combined estimates, as well as individual model estimates, were discussed at a weekly meeting between the UKHSA Epi-Ensemble and SPI-M modellers, wider SPI-M-O members and wider representatives from UKHSA and DAs. These meetings gave the modellers the chance to explain their outputs, discuss the model behaviour and agree on the inclusion or exclusion of any specific models in the ensemble for that week. A model would only be excluded if there was a clear error in its outputs, or if it displayed behaviour which could not be justified from an epidemiological perspective. Once a consensus was reached for each of the epidemic metrics, a recommendation was made to the EMRG, who then finally approved and published the consensus outputs.

### 2.4 Sensitivity analysis

Two sensitivity analyses explored the extent to which the combined *R* would have been impacted by variable weighting of the models within the ensemble and the size of the ensemble. For consistency, no individual models were re-run for these analyses; we used only the original model results submitted at the time the consensus *R* was published. This was intentional so that the analysis would serve as a historic record of the combined estimates at the time.

#### 2.4.1 Exploring the impact of model weighting on the combined *R*

Firstly we explored the impact of the choice of model weighting on the consensus *R*. The combined estimate *y* was calculated from the true effect size of each model *y*_*i*_. The true effect size can therefore be weighted. The simplest method is that of equal weighting, which was used to generate the published consensus *R* over 2020-2022. In this method, each model is assumed to have an equal contribution to the combined estimate under the assumption that all models are equally valid.

Another common method of weighting is that of inverse-variance weighting. In this method, models with a high variance, i.e. those that are less certain, are penalised more than models with a low variance, i.e. more certain models. However, individual models have different methods of representing uncertainty, and a model that is more certain is not necessarily more likely to be accurate. Therefore this method is not applicable here.

An alternative method of model weighting is to group models by either their structure, or by the data to which they fit. For example, models that may have a different structure but use the same data form a sub-group as described in Table 1. We explored the impact of this on the consensus *R* value by dividing the ensemble into sub-groups, so each sub-group represents a homogeneous set of models either according to structure or to the data to which they fit. Models within each sub-group were equally weighted, and then the contributions from the sub-groups were equally weighted to give the overall combined estimate. This had two purposes: firstly, a single data-stream or model structure would not have gained a larger weighting in the final combination, meaning that the combination was ‘data-agnostic’ or ‘model-agnostic’ and models such as EpiEstim, with a larger representation in the ensemble, did not bias the final estimates; secondly, it allowed us to compare the difference in trends between admissions and case data and therefore learn about the epidemic dynamics by inspection.

Similarly as for the equal weighting models method, a consensus *R* value was derived with this alternative variable weighting method as a range for April and September 2021. We present the results as rounded to two decimal places. However, we note that the range was published to only one decimal place to avoid presenting a false sense of precision. The range published was also rounded out, rather than rounding to the nearest decimal place, in order to increase the uncertainty instead of possibly reducing it.

#### 2.4.2 Exploring the impact of ensemble size on the combined *R*

The models included within the ensemble varied throughout the pandemic; as new models were developed and introduced, some were phased out and others were updated in response to the changing epidemic. This could hypothetically result in inconsistent estimates through time. Furthermore, as UKHSA moved from a ‘response’ to a ‘business-as-usual’ phase during 2022, a need emerged to reduce the resource dedicated to modelling COVID-19, and hence reduce the number of models in the ensemble. These factors motivated us to explore how the combined *R* may have changed if a different model ensemble was used to generate it.

We investigated the implications of reducing the size of the ensemble on the combined *R* estimate over the period April 2021 - December 2021. UKHSA models are labelled in Table 1 and comprise internal models i.e. run by UKHSA or DA modellers. We re-calculated the combined estimate using the “reduced” ensemble of only internal models and, using equal weighting, compared this to the published consensus *R* number in England.

#### 2.5 *R* as an epidemic indicator

The *R* time series is a transform of epidemic metrics such as cases incidence or hospitalisations. Hence, we expect it would be statistically correlated to the epidemic metrics, but quantifying the degree of correlation with different metrics is interesting.

We explored the correlation between the consensus *R* as published on the UK government COVID-19 dashboard and the key public data sources relating to the COVID-19 pandemic; namely, cases, admissions and deaths. We expect the *R* number to be correlated in some way to the rate of change of these three metrics, and we know this relationship is non-linear, and hence used Spearman’s rank correlation coefficient, *ρ*.

In order to adjust for weekly seasonality, each source of data was transformed to a centered weekly moving average. For each date that an *R* number was calculated, the slope of the data was calculated over a centered weekly window. We used the same length and position of windows over which to perform the analysis in order to ensure consistency, otherwise additional artificial lag would be introduced into the analysis.

The correlation between *R* and the weekly rate of change in cases, admissions or deaths may have an inherent lag due to the fact that it takes time for more severe symptoms to develop. In order to investigate this, we explored how the correlation changed between the *R* number, shifted along its time axis by a varying number of days, and the rate of change of new hospital admissions and deaths. This was done by shifting the calculated values of *R* we used by 1 - 20 days and observing how *ρ* changed with an increasing shift size. Mathematically, we are calculating the following, where the variable *X* represents the centred weekly rolling average of either the recorded incidence of cases, hospital admissions or deaths:

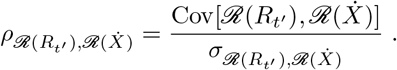

In the above, *ℛ* (·) denotes the ordinal rank and *R*_*t′*_ is the time shifted *R*, equal to *R*(*t* − *t*_shift_) where *t*_shift_ ∈ (1, 20). 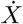 denotes the rate of change of variable *X* with respect to time, and all times considered are measured in days. We performed this calculation on data within specific time windows, which correspond to the Delta and Omicron waves respectively, and shifted the time window for the published *R* value against the static recorded data. These time windows were May 7, 2021 to July 30, 2021 and November 26, 2021 to February 25, 2022 for the Delta and Omicron waves respectively.

## 3 Results

### 3.1 Generating a consensus *R* range in April and September 2021 using different weighting methods

Whisker plots of the 90th confidence intervals of *R* for each model are plotted alongside the resulting combinations from the different methods, and shown in Figure 1. The numerical values for the 90% confidence intervals for each weighting method are given in Table 2.

**Table 2.** 90% confidence intervals for combined *R* estimates using different weighting methods.

| Weighting methodology | April 6, 2021 | September 14, 2021 |
| --- | --- | --- |
| Equal weighting | $[0.81, 0.93]$ | $[0.91, 1.07]$ |
| Weighting by data | $[0.82, 0.93]$ | $[0.86, 1.04]$ |
| Weighting by model | $[0.80, 0.94]$ | $[0.90, 1.07]$ |
| Equal weighting of case models | $[0.81, 0.90]$ | $[0.98, 1.15]$ |
| Equal weighting of hospital models | $[0.78, 1.02]$ | $[0.80, 0.93]$ |
| Equal weighting of mixed models | $[0.74, 1.00]$ | $[0.72, 1.12]$ |

**Table 3.**
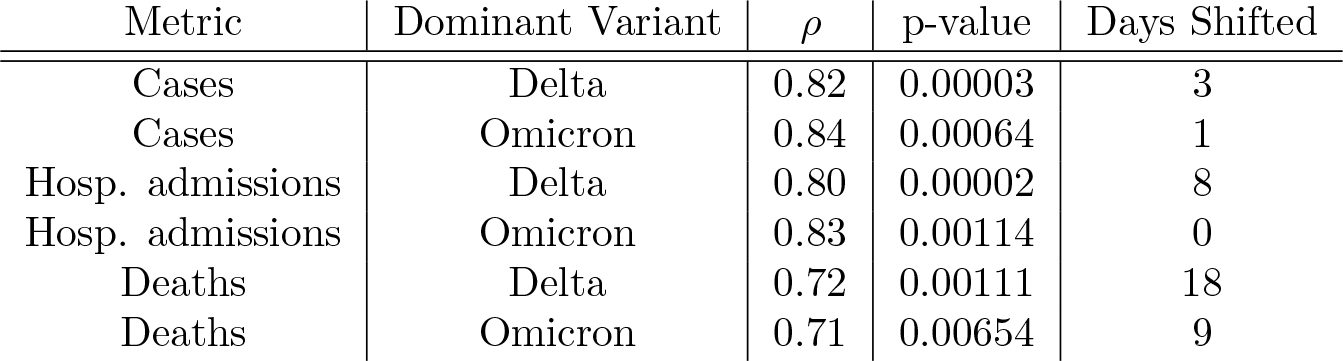
Spearman’s rank coefficient, *ρ*, and the respective p-values between the time shifted *R* and the rate of change in a given epidemic metric. The coefficient was calculated only on data within the time period shown in the table.

| Metric | Dominant Variant | $\rho$ | p-value | Days Shifted |
| --- | --- | --- | --- | --- |
| Cases | Delta | 0.82 | 0.00003 | 3 |
| Cases | Omicron | 0.84 | 0.00064 | 1 |
| Hosp. admissions | Delta | 0.80 | 0.00002 | 8 |
| Hosp. admissions | Omicron | 0.83 | 0.00114 | 0 |
| Deaths | Delta | 0.72 | 0.00111 | 18 |
| Deaths | Omicron | 0.71 | 0.00654 | 9 |

Using the equal weighting method, and combining the *R* outcomes from the various epidemiological models (a mixture of SEIR-type, agent-based and data-driven models) we generated combined *R* estimates of [0.81, 0.93] in April 2021 and [0.91, 1.07] in September 2021. These represent the 90% confidence interval that was published on the UK Government dashboard at the time.

**Figure 1.**
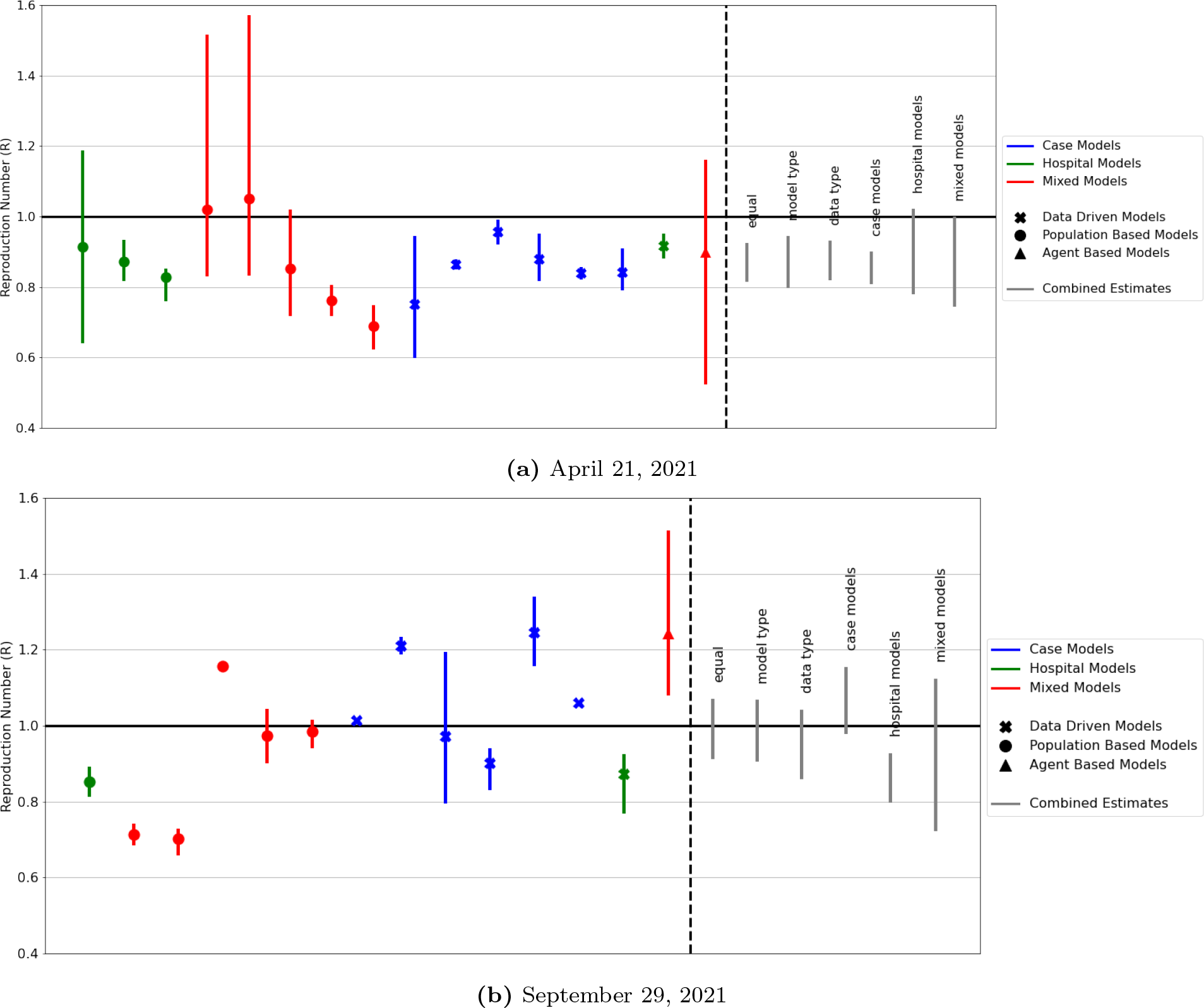
Model ensemble generated *R* values at two time points of the COVID-19 epidemic in England. The whiskers plot shows the median and the 10th and 90th percentile of the combined reproduction number *R* for the models included in the model ensemble on April 21, 2021 and September 29, 2021. The dot shows the median value while the width of the error bar for each model represents the 90% confidence interval(CI). We note that the published *R* value only showed the 90% CI. The values on the right of the dashed line show the 90% CI for the combined *R* value generated with different weighted methods. The combined *R* values are reported for April 6, 2021 and September 14, 2021.

Using different weighting for the combination of models produces very similar combined *R* values at the two snapshots in time we studied: in April 2021 and in September 2021. Weighting by data resulted in *R* combination of [0.82, 0.93] and [0.86, 1.04] for the April 2021 and September 2021 estimates respectively. Weighting by model structure resulted in a combination of [0.8, 0.94] and [0.9, 1.07] for the April 2021 and the September 2021 estimates respectively.

### 3.2 The effect of ensemble size on the combined estimate

Figure 2A compares the un-rounded combined *R* number generated from a reduced model ensemble that includes models run by UKHSA and DA teams as outlined in section 2.4.2. Our results show that the two combined *R* value time series are similar but not identical, with the level of agreement changing over the study period. For most of the study period, the values of the combined *R* from the two model ensembles were similar, with smaller model ensemble increasing the uncertainty in the consensus *R* value (comparing the width of the blue and orange bandwidth in fig. 2A). There was a notable difference in the combined *R* from the two ensembles in July 2021 which is due to a very different number of models constituting the model ensemble. The full ensemble for July 14, 2021 contained thirteen different models, compared to the internal models ensemble, which contained four (fig. 2B). On July 21, 2021 the full ensemble had eleven models, while the internal models ensemble contained only two. The two models for July 21, 2021 would not have been sufficient to produce a published combination^1^, but we have shown the result here for completeness. From August 2021 onwards the full and internal models only ensembles show much better agreement, which is due to the latter having a more comparable number of models.

**Figure 2.**
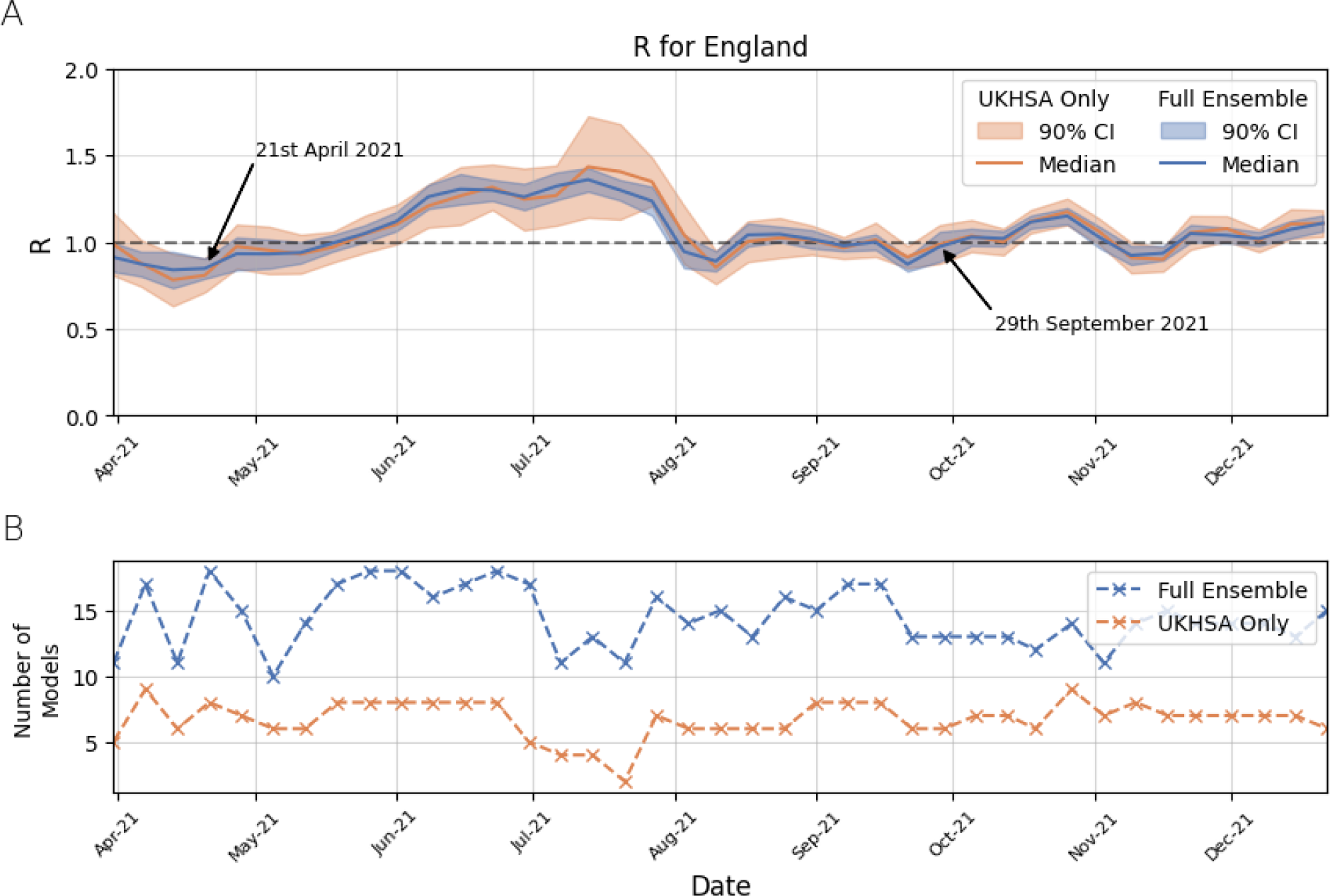
The combined *R* number in the period April 2021 - December 2021 in England for the full model ensemble and the reduced (internal UKHSA and DA models only) ensemble. Plot A shows the time series of the two *R* values over the study period, while plot B shows the number of models in each ensemble at different time points when the *R* value was generated.

### 3.3 The combined *R* is a good, but delayed, epidemic indicator

Figure 3 shows the relationship between the rate of change of the 7 day rolling mean of cases, admissions and deaths with an optimally time shifted *R*. Figure 3A shows that positive and negative *R* values (shown in red and blue respectively) occur when the number of COVID-19 cases is increasing and decreasing. The correlation, calculated as the Spearman’s rank coefficient between a time shifted *R* and the rate of change of recorded cases, is given in the box to the top left of the plot. This is done separately for the Delta and Omicron waves, and the time periods we considered for each wave are demarcated by the vertical dotted lines. Overall, our results show a good positive correlation between epidemic status indicators and a time shifted *R* across both epidemic waves, confirming that *R* is following the trends in cases, hospitalisations and deaths related to COVID-19 over both of the Delta and the Omicron epidemic waves, albeit with a delay. Here, we have shown only the maximum correlation obtained from the optimal shift of the *R* number. The values of *ρ* calculated for *R*_*t−t*shift_ where *t*_shift_ ∈ (1, 20) are shown in fig. B 1.

**Figure 3.**
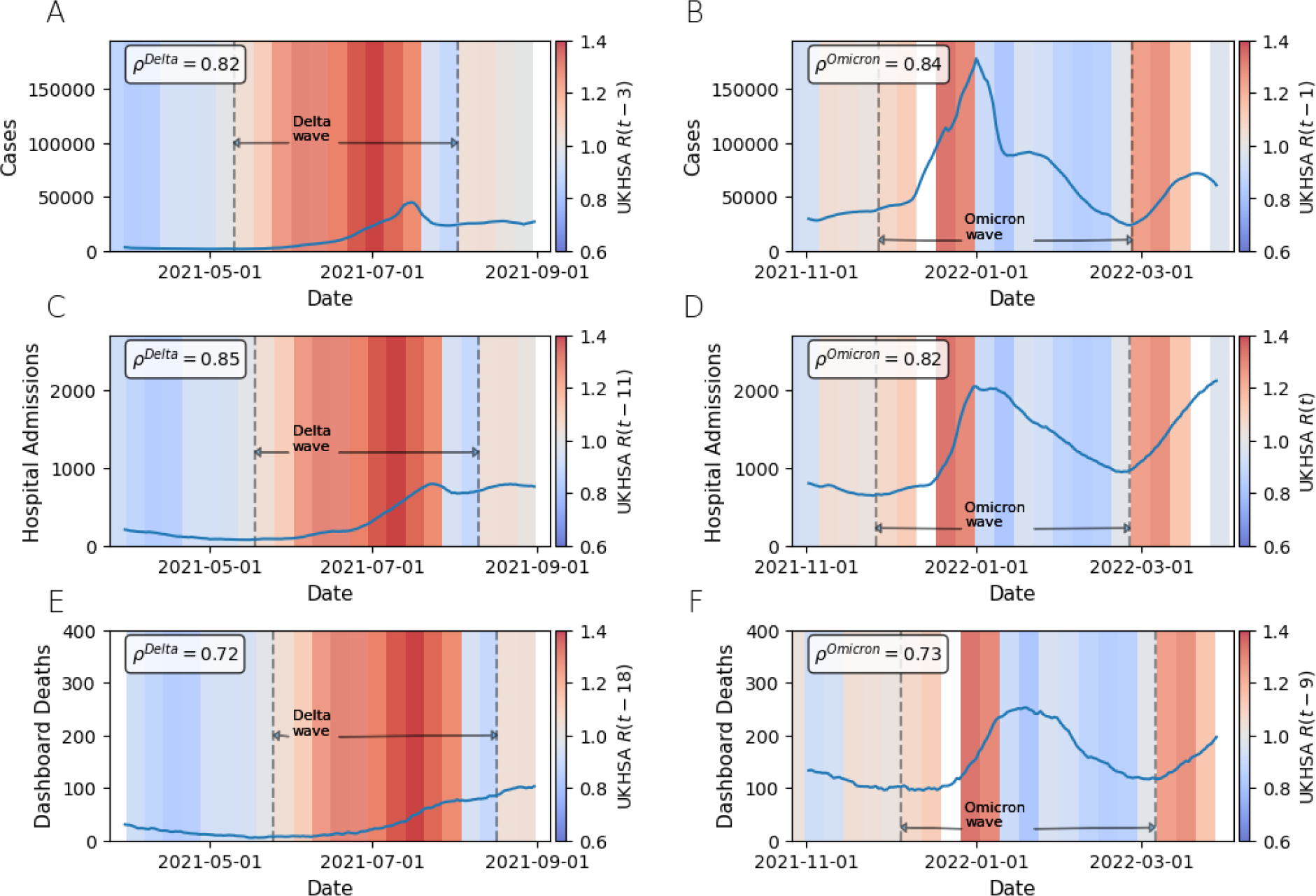
Plots comparing the published *R* number to data published on the public government COVID-19 dashboard. The plots show the superimposed time series of the 7 day rolling average of the dashboard data for various metrics, on top of the published *R* number for England. Where the shading is red, the median estimate for the *R* number was greater than 1. Where it is blue, the median *R* was less than 1. In plots C - F. For each plot, the Spearman’s rank correlation coefficient, *ρ*, was calculated to evaluate the correlation between the rate of change of the rolling 7 day mean of a given epidemic metric (cases, hospital admissions and deaths) and the median published *R* number, where *R*(*t*) has been shifted along the time axis to maximise the correlation, and *t* is measured in days. The amount of shift is different for each metric and wave. The maximum *ρ* is obtained at a shift of 3 days for the Delta wave and 1 days for the Omicron wave for cases; 9 days for the Delta wave and no shift for the Omicron wave for hospital admissions; and 18 days for the Delta wave and 9 days for the Omicron wave for deaths. Only the data within the dotted lines pertaining the Delta and Omicron waves respectively was included in the correlation calculation.

## 4 Discussion

This study outlines a collaborative approach across government and academia to generate the combined *R* value for England over the period April 2021 to December 2021 using a previously established combination method [42] and *R* estimates from an ensemble of epidemiological models. The combined *R* value was used to track the epidemic status over the COVID-19 epidemic in England, and was produced by SPI-M-O in 2020, and by the UKHSA Epi-Ensemble modelling team since early 2021.

In this paper we described the process of cross academia and government collaboration, outlined the ensemble of epidemiological models used to generate individual *R* values in England, highlighting their key structural characteristics and the data they used, as well as the method to individually derive an *R* value. We also outlined the methodology developed in [42] of combining the individual *R* values to generate a combined consensus *R* value and illustrated this by generating the published *R* values of [0.81, 0.93] on April 21, 2021 and [0.91, 1.07] on September 29, 2021.

April 21, 2021 and September 29, 2021 were very different epidemic points in time. April 21, 2021 followed the third national lockdown in England imposed to control the transmission of the Alpha variant [28]. Incidence and prevalence within the population were low and large scale vaccination against COVID-19 had only started to be rolled out, with roughly half the population having received a first dose, and only 8% having received a second dose. Against this mostly homogeneous immunity, susceptibility and vaccine backdrop, the assumptions within the models would have been similar, producing similar *R* values across models.

In September 2021, the immunity, susceptibility and vaccination levels were very different. There was a backdrop of population immunity from either vaccination or previous infection, with a large proportion of the population aged 12 and over either having received two doses of the vaccines or having been infected by the large Delta epidemic wave over the summer 2021. The COVID-19 case rate remained high with schools just returning, and this period preceded the arrival of the Omicron variant.

Different models would have made different assumptions on the impact of the large Delta wave on population immunity, and would have incorporated different assumptions around vaccination and social mixing associated with returning to school. All of these assumptions would impact individual *R* values, illustrated by the varying *R* values across models at this time.

Furthermore, different models were fit to different data and this can generate different estimates. For example, the two LSHTM EpiNow2 models, one that fits to cases, and the second that fits to admissions, have vastly different *R* estimates. This difference is also reflected in the combinations from models that fit only to cases (reporting a range of [0.98, 1.15]) and from models that fit only to hospital data (reporting a range of [0.8, 0.93]). If we were only to use models that fit to cases, this would imply that the epidemic was increasing. However, models that fit to hospital data imply that the epidemic was decreasing. Models that fit to both report a central estimate in between the two with larger uncertainty. A more thorough study of different weighting methods and their effects on the combination estimate is out of the scope of this paper, however, this relatively simple example demonstrates that it is important that the ensemble features models that fit to a range of different data sources.

We note that the ensemble of models on April 21, 2021 and September 29, 2021 are not identical, and the model ensemble has been changing over time. New models were introduced to the ensemble throughout the epidemic, models were omitted from or not submitted to the ensemble due to technical issues, such as calibration error or computer outage. Furthermore, in periods of change, such as the introduction of a new variant, some models had required extensive development work before re-inclusion into the ensemble. This is an inevitable part of the process when collaborating with various modelling teams across government and academia, who are responsively modelling a fast changing epidemic.

Reducing the size of the model ensemble to include only models run internally within UKHSA and DAs made small difference to the combined *R* value, but did increase the width of the 90% confidence interval. Overall, and for the majority of the study period, the values of the combined *R* from the two model ensembles were similar as shown in Figure 2A. There were some differences around the peaks of the Delta epidemic waves in the summer 2021, when the internal models ensemble (comprising UKHSA and DAs only models) had a very small number of models (Figure 2B) and as a consequence the combined *R* had a wider confidence interval. This suggests that our process was robust to changes in the model ensemble, provided there were more than five constituent models going into any combination. This is encouraging for institutions that may be nowcasting future epidemic: an ensemble does not need to be enormous to reap the benefits of model combination.

The time series of the combined *R* for the duration of the Delta and Omicron waves respectively are strongly positively and statistically significantly correlated to the rate of change of cases, hospitalisation and deaths related to COVID-19 (Figure 3). However, this strong positive correlation only occured for each metric if the time series for *R* was shifted along it’s time axis by a certain optimum number of days which differs for each wave and metric. Exactly what causes the specific lag for each wave and metric is unclear. We acknowledge the limitations of using the Spearman’s rank correlation coefficient to show this relationship, however, for this paper we simply wanted to gain an understanding of whether or not the *R* number is a valid proxy for epidemic status. Therefore a more sophisticated regression model, combined with a full investigation of the cause of the lag between epidemic metrics and *R*, is left to future work.

In order to mitigate uncertainty associated with nowcasting, since March 2021 the *R* value from each model was taken on a single day in time 2 weeks before the day on which models were combined^2^. Incorporating these delays in *R* is important as not all models are always able to report estimates up to the day that they are run as they do not possess the ability to forecast. For example, the simplest model, an application of *EpiEstim*, uses a delay distribution between infection and the observation to which it is fit, to back-calculate and infer the incidence time series. The *R* number is then estimated directly from the back-calculated time series for incidence. Therefore, the model is only able to provide estimates lagged to the order of the length of the delay distribution. Even where models are able to estimate current *R* numbers, due to the delay between infection and observation, the infections occurring on a given day correspond to data that will be observed in the future, and hence, are in essence, projections. Due to the difficulty in producing accurate estimates for *R* without the time delay, and in light of the above discussion about the lagged correlation, it is vital to use a range of metrics to inform policy decisions around epidemic status. For this reason the *R* value estimates were used alongside estimates of 3 other epidemiological metrics when informing policy decisions: the growth rate, *r*, incidence and prevalence, and the medium term projections for hospital admissions, occupancy and deaths.

### 4.1 Future planning and lessons learnt

While combining multiple models, particularly in epidemic modelling, has proven to be very useful during the COVID-19 epidemic, there are lessons from this that should be considered in future.

Firstly, it should be ensured that confidence intervals calculated by each of the models, represent the same sources of uncertainty. Do they capture the underlying uncertainty present in the data, the parametric uncertainty or the structural uncertainty? The forecast hub at the CDC treats models primarily as black boxes, though model details are published and models are assessed for accuracy, and there is not explicit treatment of the resulting uncertainty. For future pandemics, there should be a clear definition of uncertainty and what it should represent.

Secondly, the combination method used to generate a consensus *R* is insensitive to the performance of individual models. Whereas for forecasts, model performance can be calculated by comparing model estimates with observed data, the *R* number is a latent variable and therefore is not observed. We rely on the expertise of modellers to ensure that models fit well to the data and make sound assumptions. In the future, developing an unbiased scoring method for individual models would help in ensuring the robustness and reliability of the individual models before combining them into an ensemble.

Finally, running an ensemble of models is resource intensive and relies on a significant amount of external expertise. If models are not to be treated as black boxes specialist expertise of academic groups continues to be required, and developing formal cross government and academia modelling hubs is necessary for ongoing cross-institutional collaboration.

## Data statement

The weekly published reproduction number *R* values data is publicly available at $https://www.gov.uk/guidance/the-r-value-and-growth-rate$. Individual model *R* values can be requested from the corresponding author, but restrictions apply to the availability of these data, which were used under license for the current study, and so are not publicly available.

## Code availability

The numerical code used in this analysis can be requested from the corresponding author. Restrictions apply to the availability of the model combining code, which was under collaborative license for the current study, and hence is not publicly available.

## Data Availability

All data used and produced in this study are available upon reasonable request to the authors.

https://coronavirus.data.gov.uk

## Acknowledgements

Estimates for *R* have been provided and combined as part of the UK wide respond to COVID-We would like to thank members of the SPI-M-O modelling group named as co-authors for continuing to provide estimates. In addition, we also acknowledge Yee-Whye Teh, Robert Hinch and Christophe Fraser (University of Oxford), Michelle Kendall and Louise Dyson (University of Warwick), Axel Gandy and Neil Ferguson (Imperial College London), Robert Moore, Conor Rosato and Simon Maskell (University of Liverpool), Leon Danon and Ellen Brooks-Pollock (University of Bristol), Sam Abbott and John Edmunds (London School of Hygiene and Tropical Medicine), Robert Shaw, Ewan Wakeman, Nicholas Groves-Kirkby and Seema Patel (NHS England) and Ross Burton (Cardiff University) for insightful ongoing discussions and input to this modelling work. We would in particular like to thank the SPI-M-O secretariat, and Graham Medley (LSHTM and SPI-M-O chair) for their support and continued discussions while conducting this analysis and while producing consensus *R* estimates.

## A Models used in the ensemble

**Table A1.** Summary of the epidemiological models used to generate *R* outcomes for the English COVID-19 epidemic. We list the names of the models, their main modelling characteristics and the data to which they are calibrated against and the method to calculate *R*.

| Model Name | Description | R estimation |
| --- | --- | --- |
| Lancaster Spatial Stochastic | A Bayesian spatial stochastic compartmental model which is fit to cases. Census commuter flow data is also used to infer mixing between Local Authorities. | $R$ is calculated as the dominant eigenvalue of the NGM. |
| Edinburgh WSS Model [4] | The WSS (Weight-Shift-Scale) model fits to case data to derive $R$ but accounts for systematic reporting errors (e.g. false positives and negatives and under-reporting). Case counts are weighted, scaled and shifted to account for the change in the size in future compartments, the delay between infection and case reporting, and to account for seasonality. | The estimated $R$ is assumed to be the combination of the true $R$ plus a stochastic term and is calculated from the rate of change in reported cases, scaled by the time lag between infection and the time of case report. |
| Manchester Model (DetSEIRwithNB) [46, 11] | A deterministic compartmental ODE model that fits to hospital admissions, hospital occupancy, ICU occupancy and deaths in hospital. $\beta$, the transmission rate, varies step-wise between change points. Change points, such as policy or behavioural changes (e.g. schools returning and lockdowns) are defined by the modeller and are used to represent changes in the epidemic. | $R$ is estimated from the most recent $\beta$ in the calibrated model. |
| University of Liverpool Model [44] | A Bayesian statistical model that comprises a deterministic compartmental transmission model governed by a system of ODEs and a stochastic observation model. Fitted to deaths, hospital admissions, and symptomatic report data from NHS 111 online. | $R$ is calculated from estimates of the daily number of infections, the infectious population, and the mean time for which individuals are infectious. |
| PHE/Cambridge Model [8, 7] | A deterministic age-structured compartmental model fitted to serology data. Google mobility data is also used. Different versions of the model have been run throughout the pandemic that fit to slightly different data streams. Two versions of the model are presented in Figure 1. Deaths/ons fits to ONS infection survey data, whereas regional/age does not. More recently during the pandemic, the regional/age model has been replaced with the admissions/ons model, which has the same model structure as deaths/ons, but fits to admissions. | $R$ is calculated as the dominant eigenvalue of the NGM. |
| Warwick Model [37] | A deterministic age-structured compartmental ODE model fitted to hospital and ICU admissions, and COVID-19 positivity rate data. | $R$ is calculated as the dominant eigenvalue of the NGM. |
| Imperial Stochastic Compartmental (sircovid) [6] | Compartmental transmission model described by stochastic difference equations fitted to deaths, hospital admissions and prevalence, tested cases in hospital beds, ICU prevalence and serology data. | $R$ is calculated as the dominant eigenvalue of the NGM. |
| EpiEstim [14, 38, 39] | EpiEstim applies the renewal equation given a time series of incidence. The implementation described in [39] is used (code available at [38]). Estimates are back-calculated from an observation, e.g. cases, to time of infection, using an assumed delay distribution. | $R$ is calculated using the renewal equation Equation (2). |
| GenomicSurveillance [58] | A spatio-temporal hierarchical Bayesian model, which fits to daily new cases and COVID-19 lineage counts. | $R$ is calculated based on the derivative of cubic-spline function fitted to the incidence. |
| Epidemia [52] | A hierarchical semi-mechanistic Bayesian model based on the renewal equation. Multiple data streams can be fit to simultaneously. Two separate versions of the model are run, one that fits to weekly admissions, and one that fits to weekly cases. The admissions version of the model was developed later in the epidemic in response to changes in the case ascertainment rate. | $R$ is specified to vary weekly according to a random walk and is calculated using the renewal equation Equation (2). |
| LSHTM EpiNow2 [2, 3] | EpiNow2 uses the renewal equation to estimate $R$, where initial infections are estimated based on the initial number of cases or hospital admissions. The relationship between cases (or hospital admissions) and infections is obtained from a convolution of the relevant delay distributions (an uncertain incubation period and reporting delay). Similarly to Epidemia, versions fitting to cases and admissions are run. | $R$ is derived using the renewal equation Equation (2). |
| LSHTM ONS inc2prev [1] | A Gaussian process model that uses PCR positivity rates published by the ONS to model incidence by convolution with the curve estimating the evolution of the probability of a positive test since time of infection. | $R$ is derived using the renewal equation Equation (2). |
| Oxford CSML Model. Dashboard on [55] | Hierarchical semi-mechanistic Bayesian model fitted to cases, similar to Epidemia and as described in [24], but with a spatio-temporal component. | $R$ is derived using the renewal equation Equation (2). |
| OpenABM [35] | Stochastic agent-based model calibrated to hospital admissions, ICU bed occupancy, deaths and vaccinations. Social mixing, Test-Trace-Isolate interventions, different SARS-CoV-2 variants and progressive vaccination are explicitly modelled. | $R$ is calculated by directly counting the number of secondary infections that are caused by each primary infection. |
| Covasim [40] | A stochastic agent-based model calibrated to COVID-19 diagnoses, hospital admissions and deaths related to COVID-19 and modelling progressive vaccine roll-outs. Social distancing and Test-Trace-Isolate interventions are also modelled and progressive SARS-CoV-2 variants are incorporated. | $R$ is calculated by directly counting the number of secondary infections that are caused by each primary infection. |

## B Hospital Admissions and Deaths with a shifted *R*

**Figure B 1.**
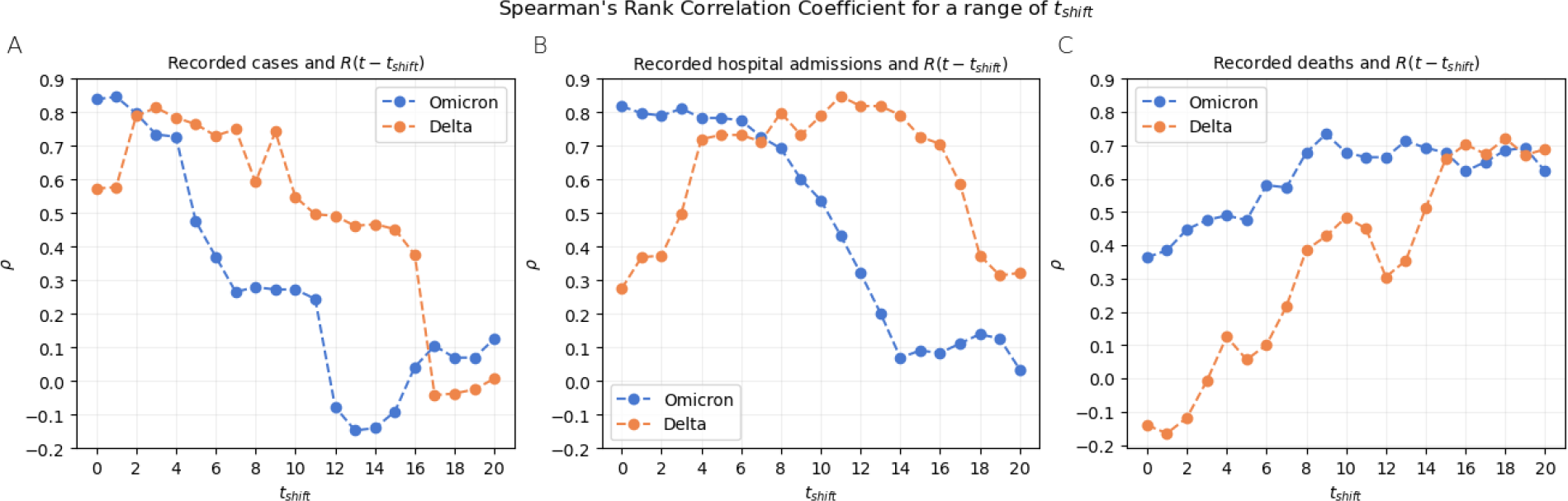
Plots A, B and C show the Spearman’s rank correlation coefficient, *ρ*, between *R*(*t − t*_shift_) and the rate of change in cases, hospital admissions, and deaths respectively, for a varying *t*_shift_. The maximum value of *ρ* found from this analysis is included in fig. 3. The minimum p-values occurred in each instance for the maximum correlations, hence the p-values are not included in this plot.

## Footnotes

1 A minimum of three distinct models was required for a combination to be published.

2 Prior to March 2021, SPI-M-O combined the most recent *R* numbers for each model, but found that there was little difference in the combined estimates produced by using estimates from two weeks prior [27].

